# Prognosis and hematological findings in patients with COVID-19 in an Amazonian population of Peru

**DOI:** 10.1101/2021.01.31.21250859

**Authors:** Sebastian Iglesias-Osores, Arturo Rafael-Heredia, Eric Ricardo Rojas-Tello, Washington A. Ortiz-Uribe, Leveau-Bartra Walter Román, Leveau-Bartra Orison Armando, Alcántara-Mimbela Miguel, Lizbeth M. Córdova-Rojas, Elmer López-López, Virgilio E. Failoc-Rojas

**Affiliations:** Facultad de Ciencias Biológicas, Universidad Nacional Pedro Ruiz Gallo, Calle Juan XXIII 391, Lambayeque, Perú; Facultad de Medicina Humana de la Universidad Nacional de Ucayali, Ucayali, Perú; Hospital II Essalud Pucallpa, Perú. Jr. Dos de Mayo Nº 521, Pucallpa; Universidad Nacional de Jaén, Jr. Cuzco 250. Jaén, Cajamarca, Perú; Universidad Señor de Sipán, Pimentel, Perú; Unidad de Investigación para la Generación y Síntesis de Evidencias en Salud, Universidad San Ignacio de Loyola, Lima, Perú

**Keywords:** Coronavirus-2019, 2019-nCoV, COVID-19, and Novel Coronavirus (SARS- CoV-2/HCoV-19, SARS-CoV-2, Coronavirus, COVID-19, Laboratory findings, Prognosis, Diagnosis

## Abstract

**Objective:** This study examined the laboratory results of COVID-19 patients from a hospital in the Peruvian Amazon and their clinical prognosis.

**Methods:** An analytical cross-sectional study was carried out whose purpose was to identify the laboratory tests of patients with COVID-19 and mortality in a hospital in Ucayali, Peru during the period from March 13 to May 9, 2020, selecting a total of 127 with Covid-19. Mean and the standard deviation was described for age, leukocytes, neutrophils, platelets, RDW-SD; median and interquartile range for the variables lymphocyte, RN / L, fibrinogen, CRP, D-dimer, DHL, hematocrit, monocytes, eosinophils.

**Results:** No differences were observed in this population regarding death and sex (OR: 1.31; 95% CI 0.92 to 1.87), however, it was observed that, for each one-year increase, the probability of death increased by 4% (PR: 1.04, 95% CI 1.03 to 1.05). The IRR (Incidence Risk Ratio) analysis for the numerical variables showed results strongly associated with hematological values such as Leukocytes (scaled by 2500 units) (IRR: 1.08, 95% CI 1.03 to 1.13), neutrophils (scaled by 2500 units) (IRR: 1.08; 95% CI 1.03 to 1.13), on the contrary, it is observed that the increase of 1000 units in lymphocytes, the probability of dying decreased by 48% (IRR: 0.52; 95% CI 0.38 to 071).

**Conclusion:** Parameters such as leukocytes and neutrophils were statistically much higher in patients who died.

## INTRODUCTION

The COVID-19 pandemic is caused by a beta coronavirus initially called 2019-nCoV or occasionally HCoV-19 and definitively named SARS-CoV-2(1). This coronavirus causes an acute respiratory syndrome that was isolated for the first time in Wuhan, China, it is believed to be of zoonotic origin, that is, it was transmitted from an animal host to a human one(2). The genome of this virus is made up of positive single-stranded RNA, this can cause transmission from person to person through drops of saliva (3). This new zoonotic virus has put the world on alert, rapidly becoming a pandemic, clinical findings in patients are important in the prognosis of the patient.

The clinical characteristics of COVID-19 are very diverse but can be arranged into four categories: systemic, respiratory, gastrointestinal, and cardiovascular (4). There are a wide number of symptoms in patients with COVID-19, only six symptoms occurred in 50% of people such as cough, sore throat, fever, myalgia or arthralgia, fatigue, and headache(5,6). Of these, fever, myalgia or arthralgia, fatigue, and headache could be considered red flags (4). Loss of smell taste is a highly specific symptom of COVID-19 (7). Knowing the specific symptoms for COVID-19 does not help to have a quick diagnosis, better follow-up, and prognosis of infected patients.

The main cause of death is respiratory failure due to acute respiratory distress syndrome (8). The overall combined mortality rate from respiratory failure in COVID-19 patients is discharged; however, this varies significantly between countries (9). People younger than 65 have a very small risk of death even in the epicenters of a pandemic, and deaths in people younger than 65 without any underlying conditions are rare. (10). The prognosis of patients with COVID-19 can be affected by comorbidities such as diabetes,(11) hypertension, stroke, cancer, kidney disease, and high cholesterol (12). Risk factors should be considered to avoid rapid progression and poor prognosis of COVID-19 disease. More attention should be paid to these patients, in case of rapid deterioration.

Laboratory medicine is of great help for the early detection of SARS-CoV-2 and allows us to discriminate between patients with severe and non-severe COVID-19 (13). Findings in COVID-19 patients will help better understand key disease traits and could be used for future disease research, control, and prevention (14). COVID-19 has a significant impact on the hematopoietic system and hemostasis (15). Leukocytosis, lymphopenia, and thrombocytopenia are associated with increased severity and even death in COVID-19 cases(16). Lymphopenia can be considered as a key laboratory finding, with prognostic potential (15). A significant decrease in peripheral CD4 + and CD8 + T lymphocytes was observed (17). These findings are important to know the prognosis of patients with COVID-19.

There are still questions to be resolved about COVID-19, it is with us a short time, which is a short time to give accurate details about it, that is why it is necessary to increase research in all fields of science, to help us fight the pandemic and get back to the new normal. This study examined the laboratory results of COVID-19 patients from a hospital in the Peruvian Amazon and their clinical prognosis.

## METHODS

### Study design

An analytical cross-sectional study was carried out, the purpose of which was to identify the laboratory tests of patients with COVID-19 and mortality in a hospital in Ucayali (located in the Central Jungle, in eastern Peru).

### Population and sample

The population consisted of hospitalized patients with a confirmed diagnosis of COVID-19 in a hospital in Ucayali, Peru during the period from March 13 to May 9, 2020, selected a total of 127 people consecutively.

### Procedures

The diagnosis of COVID-19 was made based on the provisional guidelines of the World Health Organization (WHO)(18). A confirmed case of Covid-19 was defined as a positive result in the polymerase chain reaction-transcriptase (RT-PCR) assay of nasopharyngeal swab samples and rapid immunochromatographic test. The only laboratory-confirmed cases were included in the analysis. Follow-up was carried out until discharge or death.

The laboratory tests were performed within the hospital headquarters at the time of admission, these were corroborated by a pathologist. All the laboratory tests were carried out following the clinical care needs of the patient following the technical standard of the Ministry of Health of Peru. Laboratory evaluations consisted of leukocytes, neutrophils, platelets, RDW-SD, RN / L, fibrinogen, CRP, D-dimer, DHL, hematocrit, monocytes, eosinophils.

### Statistical analysis

Statistical analysis was performed in the STATA v.16.1 software (StataCorp LP, College Station, TX, USA). Mean and the standard deviation was described for age, leukocytes, neutrophils, platelets, RDW-SD; median and interquartile range for the variables lymphocyte, RN / L, fibrinogen, CRP, D-dimer, DHL, hematocrit, monocytes, eosinophils; the variables sex, ICU, and death were reported as frequencies and percentages.

In the bivariate analysis of categorical variables, the chi-square test was used to explore the association between laboratory factors and COVID-19 mortality, after evaluating the assumption of expected frequencies. For the numerical variables, the student’s t-test was used for independent samples, or the Mann Whitney U test according to the normality distribution. In the simple regression analysis, prevalence ratios and 95% confidence intervals were estimated, using the Poisson distribution family, log link function, and robust variance. For the multiple models, a parsimonious model was built by performing a nesting process between the variables that were found to contribute significantly to the model, using the Likelihood-ratio test.

The area under the curve (AUC) values with their 95% confidence intervals of the hematological variables were calculated for the prediction of mortality.

### Ethical aspects

This research has been approved by the Ethics Committee of the National University of Ucayali. Codes were used to maintain the confidentiality of data for COVID-19 patients eligible for this study. The ethical principles of the Declaration of Helsinki were respected.

## RESULTS

Information was collected from 127 people from Hospital II Essalud Pucallpa. It is a population with an average age of 59.6 years, where the female sex predominated (67.72%). Of this population, it was found that 21 (83.46 %) patients were in the ICU, also the entire population had a mortality of 75 people (59.06 %).

Regarding the laboratory tests, it is observed that, on average, the general population had high levels of biochemical and hematological values. However, when stratifying according to mortality, the parameters such as leukocytes and neutrophils were statistically much higher in patients who died (p <0.001 in both cases), also, those who died had lymphocytopenia compared to normal values in survivors (720 vs 1360 respectively, p <0.001), something similar happened with monocytes and eosinophils.

It can be seen that in other parameters such as RN / L, fibrinogen among others, they were higher in the population that died compared to the one that survived. Platelet, RDW-SD, and D-dimer values were statistically similar in both groups. See Table 1.

**Table 1:** Clinical, laboratory, and hematological characteristics of the population hospitalized with COVID-19, Ucayali.

|  | Total |  | Died |  | Survived |  |  |
| --- | --- | --- | --- | --- | --- | --- | --- |
| **Numeric variables \*** | **Mean** | **SD** | **Mean** | **SD** | **Mean** | **DS** | **p-value** |
| **Age** | 59.6 | 14.21 | 66.1 | 10.8 | 50.2 | 13.3 | <0.001 |
| **Leukocytes** | 13 778.12 | 6 973.44 | 15 503.9 | 7 438 | 11 289.1 | 5 400 | <0.001 |
| **Neutrophils** | 12 037.04 | 7 183.8 | 14 034.5 | 7 413 | 9 099.6 | 5 735 | <0.001 |
| **Platelets** | 286 119 | 130 685 | 290 202 | 14 156 | 280 307 | 19 890 | 0.677 |
| **RDW-SD** | 45.42 | 3.9 | 45.3 | 3.6 | 45.6 | 4.5 | 0.642 |
| **Numeric variables ±** | **Median** | **IQR** | **Median** | **IQR** | **Median** | **IQR** | **p-value** |
| **Lymphocyte** | 900 | 690 | 720 | 520 | 1360 | 1770 | <0.001 |
| **AT** | 12.3 | 17 | 17.26 | 21.9 | 5.56 | 11.5 | <0.001 |
| **Fibrinogen** | 672 | 490 | 774.3 | 264 | 373.5 | 182 | <0.001 |
| **PCR** | 2.45 | 13.5 | 1.8 | 2.8 | 3.85 | 15.4 | 0.012 |
| **D-dimer** | 1.48 | 2.7 | 4.7 | 6.2 | 1.35 | 1.6 | 0.091 |
| **DHL** | 715 | 390 | 817 | 261 | 495.5 | 298 | <0.001 |
| **Hematocrit** | 39.5 | 8.2 | 42.2 | 6.4 | 37.65 | 7.4 | <0.001 |
| **Monocytes** | 390 | 300 | 300 | 300 | 480 | 390 | <0.001 |
| **Eosinophils** | 20 | 80 | 10 | 45 | 125 | 290 | <0.001 |
| **Categorical variables¥** | **N** | **%** | **N** | **%** | **N** | **%** | **p-value** |
| **Sex** |  |  |  |  |  |  |  |
| **Male** | 86 | 32.28 | 20 | 48.78 | 31 | 36.05 | 0.104 |
| **Female** | 41 | 67.72 | 55 | 63.95 | 31 | 36.05 |  |
| **RN/L Categorized** |  |  |  |  |  |  |  |
| **RN/L>= 30** | 101 | 80.16 | 71 | 70.3 | 30 | 29.7 | <0.001 |
| **RNL/L <30** | 25 | 19.84 | 4 | 16 | 21 | 84 |  |
| **Condition** |  |  |  |  |  |  |  |
| **UCI** | 21 | 83.46 | 5 | 23.8 | 16 | 76.2 | <0.001 |
| **No-UCI** | 106 | 16.54 | 70 | 66.1 | 36 | 33.9 |  |

SD: Standard deviation. IQR: Interquartile range. \* Values are presented in mean and SD, p-value calculated with the t-student test. ± Median and IQR values are presented, p-value calculated with the Mann-Whitney test. ¥ Absolute and relative frequencies are presented, p-value calculated with the chi-square test.

No differences were observed in this population regarding death and sex (OR: 1.31; 95% CI 0.92 to 1.87), however, it was observed that, for each one-year increase, the probability of death increased by 4% (PR: 1.04, 95% CI 1.03 to 1.05). The IRR (Incidence Risk Ratio) analysis for the numerical variables showed results strongly associated with hematological values such as Leukocytes (scaled by 2500 units) (IRR: 1.08, 95% CI 1.03 to 1.13), neutrophils (scaled by 2500 units) (IRR: 1.08; 95% CI 1.03 to 1.13), on the contrary, it is observed that the increase of 1000 units in lymphocytes, the probability of dying decreased by 48% (IRR: 0.52; 95% CI 0.38 to 071). It is worth mentioning that the one with the greatest strength of association was the D-dimer since for each 1 mg / L increase in D-dimer, the risk of dying increased by 25% (IRR: 1.25, 95% CI 1.10 to 1.43). The other results are shown in Table 2.

**Table 2:** Comparison of hematological and clinical values in the hospitalized population with COVID-19, Ucayali.

|  | Crude Analysis |  |  | Parsimononic model |  |  |
| --- | --- | --- | --- | --- | --- | --- |
|  | RP | CI 95% | p* | RP | CI 95% | p± |
| **Sex** |  |  |  |  |  |  |
| **Female** | Ref |  |  | - |  |  |
| **Male** | 1.31 | 0.92 a 1.87 | 0.133 |  |  |  |
|  | **IRR** | **IC 95%** | **valor-p** | **IRR** | **IC 95%** | **valor-p** |
| **Age** | 1.04 | 1.03 a 1.05 | <0.001 | - |  |  |
| **Neutrophils (Staggered 2500)** | 1.09 | 1.05 a 1.14 | <0.001 | 1.08 | 0.99 a 1.17 | 0.089 |
| **Lymphocyte (Staggered 1000)** | 0.52 | 0.38 a 0.71 | <0.001 | 0.22 | 0.08 a 0.57 | 0.002 |
| **D-dimer** | 1.25 | 1.10 a 1.43 | 0.001 | 1.14 | 1.01 a 1.29 | 0.039 |
| **Hematocrit** | 1.04 | 1.01 a 1.07 | 0.008 | - |  |  |
| **RDW-SD** | 0.99 | 0.95 a 1.03 | 0.652 | - |  |  |
| **Platelets (Staggered 1000000)** | 1.27 | 0.41 a 3.97 | 0.682 | - |  |  |
| **Leukocytes (Step 2500)** | 1.08 | 1.04 a 1.13 | <0.001 | - |  |  |
| **RN/L** | 1.004 | 1.00 a 1.0007 | 0.020 | - |  |  |
| **Fibrinogen (Tier 100)** | 1.03 | 1.01 a 1.06 | 0.013 | - |  |  |
| **PCR** | 0.98 | 0.085 a 1.01 | 0.085 | - |  |  |
| **DHL** | 1.00 | 1.00 a 1.00 | 0.340 | - |  |  |
| **Monocytes** | 0.99 | 0.99 a 1.00 | 0.226 | - |  |  |
| **Eosinophils** | 0.99 | 0.99 a 1.00 | 0.053 | - |  |  |

\*P-value obtained from the generalized linear model, using the Poisson family and the robust Log and variance link function. ± p-value obtained from the generalized linear model for a parsimonious model with backward elimination, Poisson family, and robust Log link function and variance were used. 95% CI: 95% confidence interval. RP: Prevalence ratio. IRR: Incidence Risk Ratio.

In the explanatory variables model, it was observed that D-dimer and lymphocytes (scaled by 1000 units) had a strong association with mortality (IRR: 1.18 and 0.22 respectively), neutrophils (scaled by 2500 units) had a marginal association (IRR: 1.08, 95% CI: 0.99 to 1.17). See Table 2.

These results were complemented with the search for the area under the curve (AUC), where it was found that fibrinogen had the best AUC (AUC: 82.5%, 95% CI 72.17 to 92.83), followed by RN / L with AUC: 79.01 %. These and other variables can be seen in table 3 and figure 1.

**Table 3:** Areas under the curve (AUC) of hematological variables and mortality prediction in patients with COVID-19 in Ucayali.

|  | **AUC (%)** | **CI 95%** |
| --- | --- | --- |
| **Leukocytes** | 67.35 | 57.94 to 76.76 |
| **Neutrophils** | 70.78 | 61.61 to 79.97 |
| **Platelets** | 53.47 | 43.03 to 63.91 |
| **RDW-SD** | 50.51 | 39.73 to 61.30 |
| **Lymphocyte** | 75.46 | 66.44 to 84.49 |
| **EN / L** | 79.01 | 70.87 to 87.15 |
| **Fibrinogen** | 82.50 | 72.17 to 92.83 |
| **PCR** | 64.39 | 53.65 to 75.14 |
| **D-dimer** | 65.14 | 44.44 to 85.83 |
| **DHL** | 73.42 | 61.33 to 85.51 |
| **Hematocrit** | 71.36 | 61.99 to 80.72 |
| **Monocytes** | 66.17 | 56.56 to 75.78 |
| **Eosinophils** | 76.59 | 64.86 to 88.33 |

RNL: Neutrophil Lymphocyte Ratio. \* AUC: Area under the curve.

**Figure 1:**
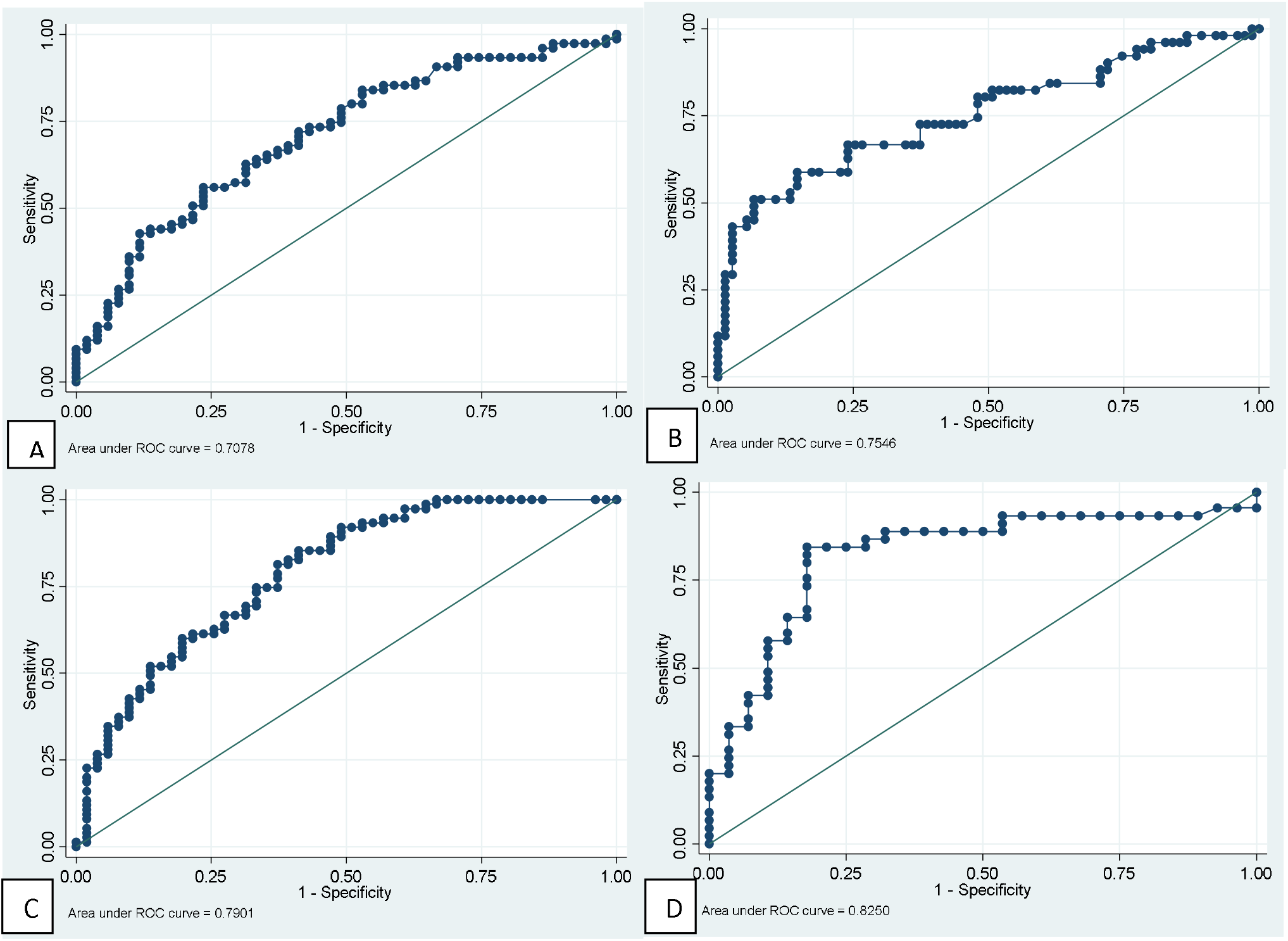
ROC curves of main hematological tests for predicting mortality in patients with COVID-19. A: Neutrophil. B: Lymphocytes. C: Neutrophil / lymphocyte ratio. D: Fibrinogen.

## DISCUSSION

We look at the clinical symptoms of 127 COVID-19 patients, here, we mainly analyze and summarize the laboratory tests of COVID-19 patients, especially the biochemical and hematological parameters. In our study, the laboratory tests of the general population studied that was infected by SARS-CoV-2 had high levels of biochemical and hematological values, these findings have also been found in other systematic reviews of patients with COVID-19 and SARS(16,19,20). These findings can be explained why the blood presents a state of hypercoagulation, which becomes more acute in critically ill patients. (21). Our results show important biochemical and hematological changes in patients with COVID-19.

It was found that the parameters such as leukocytes and neutrophils were statistically much higher in patients who died (p <0.001 in both cases), these risk factors related to immunity can predict the mortality of patients with COVID-19 (22). Patients who died presented lymphocytopenia compared to normal values in survivors (720 vs 1360 respectively, p <0.001), something similar happened with monocytes and eosinophils. Lymphocytopenia is an independent risk factor for hospital mortality for COVID-19 patients, especially men(23). Risk factors related to the mortality of patients with COVID-19 were found such as increased leukocytes and neutrophils along with lymphocytopenia, the evaluation of these parameters can help to identify people at high risk with COVID-19.

It can be seen that in other parameters such as the neutrophil-lymphocyte ratio (RN / L), fibrinogen among others, they were higher in the population that died compared to the one that survived. Elevated fibrinogen level has been considered an epiphenomenon of pulmonary edema rather than activation of coagulation in affected patients(24). Platelet values, red cell distribution width with standard deviation (RDW-SD), and D-dimer were statistically similar in both groups. There are findings in which an increase in D-dimer levels is found and this increases according to the severity in patients with COVID-19 (24,25). RDW-SD was a predictor in the group of severe patients in which its values were significantly higher than those of the group of patients with moderate COVID-19 (26).

In this study, it was observed that, for each one-year increase, the probability of dying increased by 4%, these results have been reported in other studies (27). It is worth mentioning that the one who presented the greatest strength of association was D-dimer, since for every 1 mg / L increase in D-dimer, the risk of dying increased by 25%, these high values are associated in those with severe disease(28).

We conclude that the laboratory tests, it is observed that, on average, the general population had high levels of biochemical and hematological values. Parameters such as leukocytes and neutrophils were statistically much higher in patients who died.

## Data Availability

The information is clinical data that should not be disclosed.

https://doi.org/10.6084/m9.figshare.13670275.v2

## References

1. Yang J, Niu P, Chen L, Wang L, Zhao L, Huang B, et al. Genetic tracing of HCoV-19 for the re-emerging outbreak of COVID-19 in Beijing, China. Protein Cell [Internet]. 2020 Aug 17 [cited 2020 Nov 11];1–3. Available from: https://doi.org/10.1007/s13238-020-00772-0

2. Zhou P, Yang X Lou, Wang XG, Hu B, Zhang L, Zhang W, et al. A pneumonia outbreak associated with a new coronavirus of probable bat origin. Nature. 2020 Mar 12;579(7798):270–3.

3. Nagura-Ikeda M, Imai K, Tabata S, Miyoshi K, Murahara N, Mizuno T, et al. Clinical evaluation of self-collected saliva by quantitative reverse transcription-PCR (RT-qPCR), Direct RT-qPCR, reverse transcription-loop-mediated isothermal amplification, and a rapid antigen test to diagnose COVID-19. J Clin Microbiol [Internet]. 2020 Sep 1 [cited 2020 Nov 11];58(9). Available from: https://doi.org/10.1128/JCM

4. Struyf T, Deeks JJ, Dinnes J, Takwoingi Y, Davenport C, Leeflang MMG, et al. Signs and symptoms to determine if a patient presenting in primary care or hospital outpatient settings has COVID-19 disease. Cochrane Database Syst Rev [Internet]. 2020 Jul 7 [cited 2020 Nov 11];2020(7). Available from: http://doi.wiley.com/10.1002/14651858.CD013665

5. Halboub E, Al-Maweri SA, Alanazi RH, Qaid NM, Abdulrab S. Orofacial manifestations of COVID-19: a brief review of the published literature. Braz Oral Res [Internet]. 2020 [cited 2020 Nov 11];34. Available from: https://doi.org/10.1590/1807-3107bor-2020.vol34.0124

6. Pan L, Mu M, Yang P, Sun Y, Wang R, Yan J, et al. Clinical characteristics of COVID-19 patients with digestive symptoms in Hubei, China: A descriptive, cross-sectional, multicenter study. Am J Gastroenterol [Internet]. 2020 May 1 [cited 2020 Nov 11];115(5):766–73. Available from: /pmc/articles/PMC7172492/?report=abstract

7. Makaronidis J, Mok J, Balogun N, Magee CG, Omar RZ, Carnemolla A, et al. Seroprevalence of SARS-CoV-2 antibodies in people with an acute loss in their sense of smell and/or taste in a community-based population in London, UK: An observational cohort study. PLoS Med [Internet]. 2020 Oct 1 [cited 2020 Nov 11];17(10):e1003358. Available from: https://doi.org/10.1371/journal.pmed.1003358

8. Ruan Q, Yang K, Wang W, Jiang L, Song J. Clinical predictors of mortality due to COVID-19 based on an analysis of data of 150 patients from Wuhan, China [Internet]. Vol. 46, Intensive Care Medicine. Springer; 2020 [cited 2020 Nov 11]. p. 846–8. Available from: https://www.ncbi.nlm.nih.gov/pmc/articles/PMC7080116/

9. Ioannidis JPA, Axfors C, Contopoulos-Ioannidis DG. Population-level COVID-19 mortality risk for non-elderly individuals overall and for non-elderly individuals without underlying diseases in pandemic epicenters. Environ Res [Internet]. 2020 Sep 1 [cited 2020 Nov 11];188:109890. Available from: /pmc/articles/PMC7327471/?report=abstract

10. Hasan SS, Capstick T, Ahmed R, Kow CS, Mazhar F, Merchant H a., et al. Mortality in COVID-19 patients with acute respiratory distress syndrome and corticosteroids use: a systematic review and meta-analysis. Expert Rev Respir Med [Internet]. 2020 Nov 1 [cited 2020 Nov 11];14(11):1149–63. Available from: https://www.tandfonline.com/doi/full/10.1080/17476348.2020.1804365

11. Guo W, Li M, Dong Y, Zhou H, Zhang Z, Tian C, et al. Diabetes is a risk factor for the progression and prognosis of COVID-19. Diabetes Metab Res Rev [Internet]. 2020 Oct 1 [cited 2020 Nov 11];36(7). Available from: https://onlinelibrary.wiley.com/doi/full/10.1002/dmrr.3319

12. Zaki N, Alashwal H, Ibrahim S. Association of hypertension, diabetes, stroke, cancer, kidney disease, and high-cholesterol with COVID-19 disease severity and fatality: A systematic review. Diabetes Metab Syndr Clin Res Rev. 2020 Sep 1;14(5):1133–42.

13. Pourbagheri-Sigaroodi A, Bashash D, Fateh F, Abolghasemi H. Laboratory findings in COVID-19 diagnosis and prognosis. Vol. 510, Clinica Chimica Acta. Elsevier B.V.; 2020. p. 475–82.

14. Xie Y, Wang Z, Liao H, Marley G, Wu D, Tang W. Epidemiologic, clinical, and laboratory findings of the COVID-19 in the current pandemic: Systematic review and meta-analysis [Internet]. Vol. 20, BMC Infectious Diseases. BioMed Central Ltd; 2020 [cited 2020 Nov 11]. p. 640. Available from: https://bmcinfectdis.biomedcentral.com/articles/10.1186/s12879-020-05371-2

15. Terpos E, Ntanasis-Stathopoulos I, Elalamy I, Kastritis E, Sergentanis TN, Politou M, et al. Hematological findings and complications of COVID-19. Am J Hematol [Internet]. 2020 Jul 1 [cited 2020 Nov 11];95(7):834–47. Available from: https://onlinelibrary.wiley.com/doi/full/10.1002/ajh.25829

16. Henry BM, De Oliveira MHS, Benoit S, Plebani M, Lippi G. Hematologic, biochemical and immune biomarker abnormalities associated with severe illness and mortality in coronavirus disease 2019 (COVID-19): A meta-analysis. Clin Chem Lab Med [Internet]. 2020 Jun 25 [cited 2020 Nov 11];58(7):1021–8. Available from: https://pubmed.ncbi.nlm.nih.gov/32286245/

17. Liu X, Zhang R, He G. Hematological findings in coronavirus disease 2019: indications of progression of disease [Internet]. Vol. 99, Annals of Hematology. Springer; 2020 [cited 2020 Nov 11]. p. 1421–8. Available from: https://doi.org/10.1007/s00277-020-04103-5

18. Organization WH. Clinical management of severe acute respiratory infection (SARI) when COVID-19 disease is suspected: interim guidance, 13 March 2020. World Health Organization; 2020.

19. Elshazli RM, Toraih EA, Elgaml A, El-Mowafy M, El-Mesery M, Amin MN, et al. Diagnostic and prognostic value of hematological and immunological markers in COVID-19 infection: A meta-analysis of 6320 patients. Afrin F, editor. PLoS One [Internet]. 2020 Aug 21 [cited 2020 Nov 15];15(8 August):e0238160. Available from: https://dx.plos.org/10.1371/journal.pone.0238160

20. Gu J, Korteweg C. Pathology and pathogenesis of severe acute respiratory syndrome. Am J Pathol [Internet]. 2007 [cited 2020 Nov 15];170(4):1136–47. Available from: https://pubmed.ncbi.nlm.nih.gov/17392154/

21. Yuan X, Huang W, Ye B, Chen C, Huang R, Wu F, et al. Changes of hematological and immunological parameters in COVID-19 patients. Int J Hematol [Internet]. 2020 Oct 1 [cited 2020 Nov 15];112(4):553–9. Available from: https://doi.org/10.1007/s12185-020-02930-w

22. Zhao Y, Nie HX, Hu K, Wu XJ, Zhang YT, Wang MM, et al. Abnormal immunity of non-survivors with COVID-19: Predictors for mortality. Infect Dis Poverty [Internet]. 2020 Aug 3 [cited 2020 Nov 15];9(1):108. Available from: https://idpjournal.biomedcentral.com/articles/10.1186/s40249-020-00723-1

23. Liu Y, Du X, Chen J, Jin Y, Peng L, Wang HHX, et al. Neutrophil-to-lymphocyte ratio as an independent risk factor for mortality in hospitalized patients with COVID-19. J Infect. 2020 Jul 1;81(1):e6–12.

24. Hayiroglu MI, Cinar T, Tekkesin AI. Fibrinogen and D-dimer variances and anticoagulation recommendations in Covid-19: Current literature review. Rev Assoc Med Bras [Internet]. 2020 Jun 1 [cited 2020 Nov 15];66(6):842–8. Available from: http://dx.

25. Tang N, Li D, Wang X, Sun Z. Abnormal coagulation parameters are associated with poor prognosis in patients with novel coronavirus pneumonia. J Thromb Haemost. 2020 Apr 1;18(4):844–7.

26. Wang C, Deng R, Gou L, Fu Z, Zhang X, Shao F, et al. Preliminary study to identify severe from moderate cases of COVID-19 using combined hematology parameters. Ann Transl Med [Internet]. 2020 May [cited 2020 Nov 15];8(9):593–593. Available from: /pmc/articles/PMC7290538/?report=abstract

27. Mallapaty S. The coronavirus is most deadly if you are older and male-new data reveal the risks. Nature. 2020;16–7.

28. Lippi G, Favaloro EJ. D-dimer is Associated with Severity of Coronavirus Disease 2019: A Pooled Analysis. Thromb Haemost [Internet]. 2020 May 1 [cited 2020 Dec 7];120(5):876–7. Available from: https://www.ncbi.nlm.nih.gov/pmc/articles/PMC7295300/

